# Cervical and systemic innate immunity predictors of HIV risk linked to genital herpes acquisition and time from HSV-2 seroconversion

**DOI:** 10.1101/2022.02.13.22270917

**Authors:** Y. Govender, C Morrison, P Chen, X Gao, H. Yamamoto, T. Chipato, S. Anderson, R. Barbieri, R. Salata, G. F. Doncel, R. N. Fichorova

**Author notes:** Corresponding author **Correspondence to**: Raina Fichorova, MD, PhD, Laboratory of Genital Tract Biology, Department of Obstetrics, Gynecology and Reproductive Biology, Brigham and Women’s Hospital, Harvard Medical School, 221 Longwood Ave, RF472, Boston, MA 02115, USA.

## Abstract

**Objectives:** To examine innate immunity predictors of HIV-1 acquisition as biomarkers of HSV-2 risk and biological basis for epidemiologically established HIV-1 predisposition in HSV-2 infected women.

**Methods:** We analyzed longitudinal samples from HIV-1 negative visits of 1019 women before and after HSV-2 acquisition. We measured cervical and serum biomarkers of inflammation and immune activation previously linked to HIV-1 risk. Protein levels were Box–Cox transformed and odds ratios for HSV-2 acquisition were calculated based on top quartile or below/above median levels for all HSV-2 negative visits. Bivariate analysis determined the likelihood of HSV-2 acquisition by biomarker levels preceding infection. Linear mixed-effects models evaluated if biomarkers differed by HSV-2 status defined as negative, incident, or established infections with an established infection cut-off starting at 6 months.

**Results:** In the cervical compartment, two biomarkers of HIV-1 risk (low SLPI and high BD-2) also predicted HSV-2 acquisition. In addition, HSV-2 acquisition was associated with IL-1β, IL-6, IL-8, MIP-3α, ICAM-1 and VEGF when below median levels. Systemic immunity predictors of HSV-2 acquisition were high sCD14 and IL-6, with highest odds when concomitantly increased (OR=2.23, 1.49-3.35). Concomitant systemic and mucosal predictors of HSV-2 acquisition risk included: 1) serum top quartile sCD14 with cervical low SLPI, VEGF and ICAM-1, or high BD-2; serum high IL-6 with cervical low VEGF and ICAM-1, SLPI, IL-1β and IL-6, and 3) serum low CRP with cervical high BD-2. Most cervical biomarkers were decreased after HSV-2 acquisition compared to the HSV-2 negative visits, with incident infections associated with a larger number of suppressed cervical biomarkers and lower serum IL-6 levels compared to established infections.

**Conclusions:** A combination of systemic immunoinflammatory and cervical immunosuppressed states predicts HSV-2 acquisition. A persistently suppressed innate immunity during incident infection may add to the increased HIV-1 susceptibility.

**Key Messages:**

- A combination of altered systemic and cervical immunity precedes and predicts risk of HSV-2 acquisition.
- Factors causing cervical mucosal imbalance (low SLPI and high BD-2) may predispose to both HIV-1 and HSV-2 acquisition
- In comparison to non-infected, HSV-2 infected women show suppressed cervical innate immunity
- Compared to women with established HSV-2 infection, those with incident infections within 6 months from seroconversion are more immunosuppressed both at the mucosal and peripheral level, adding to the biology of HIV predisposition.

## Introduction

The significance of herpes simplex virus-2 (HSV-2) as a risk factor for HIV is driven by the high HSV-2 prevalence worldwide, estimated to be 39.3%--83.3% in South African women^1^. Women and men infected with HSV-2 have estimated three-fold higher HIV acquisition^2^. A 3.2- and 4.6-fold increased risk of HIV acquisition was associated with HSV-2 among 4500 Ugandan and Zimbabwean women, respectively^3^. The same study found greater HIV acquisition with recent (within 3-21 months from a negative test) compared to prevalent HSV-2 infections in both Uganda (4.6-vs. 2.8-fold) and Zimbabwe (8.6-vs. 4.4-fold). These data are supported by findings of greater frequency and severity of clinically active herpes episodes after recent HSV-2 infections^2^, which cause breaches in the epithelial layer and influx of activated immune cells, providing HIV with access to target cells ^4,5^. The possibility of HSV-2-initiated clinical or subclinical mucosal inflammation and innate immunity imbalance ^5,6^ has been proposed as additional mechanism linking these viral infections but needed validation in a large clinical study.

We previously identified biomarkers of innate immunity predicting HIV-1 risk in the Hormonal Contraceptive and Risk of HIV (HC-HIV) cohort – one of the largest prospective studies examining the role of HC in HIV acquisition among African women^7,8^. Both mucosal and systemic immunity imbalances contributed to the HIV risk^7^ and aberrant cervical immunity preceded other sexually transmitted infections including HSV-2.^8^ It remained unknown whether the immunity imbalance predisposing to HSV-2 is limited to the cervical compartment or extends to the systemic circulation and whether HSV-2 in turn may alter both mucosal and peripheral innate immunity to contribute to HIV-1 risk. To address the gap in our understanding of the impact of mucosal and systemic immunity on HSV-2 acquisition and HSV-2 immunity related to HIV risk, we investigated the following hypotheses: 1) aberrant systemic immunity concomitantly with altered cervical immunity precedes and predisposes to HSV-2 infection, 2) HSV-2 infection changes cervical and systemic innate immunity, and 3) these changes may depend on the duration of HSV-2 infection which may add to the biological explanation of the greater HIV acquisition risk in incident versus established HSV-2 infections. To this end, we analyzed longitudinal cervical and serum specimens collected from participants in the HC-HIV study and defined two models for our analysis based on: 1) HSV-2 status by visit (negative, incident or established), and 2) HSV-2 status by participant (remained negative or HSV-2 seroconverted).

## Methods

### Human subject research ethics

This analysis of biospecimens from the HC-HIV study received approval from institutional review boards at FHI 360 and the Brigham and Women’s Hospital. The transfer of samples received institutional approvals from the authorities in Zimbabwe and Uganda.

### Study Population and Study Visits

Biospecimens from 5193 HIV-negative study visits from 1275 women were available from the HC-HIV study. Infections at study visits within 6 months (180 days) after first becoming HSV-2 seropositive were considered incident while those from visits >6 months after a visit with confirmed positive seroconversion were considered established. The biological rationale for choosing the 6-month cut-off was based on observations of more HSV shedding within the first 6 months after acquisition^9^ and decreased clinical reactivation over time^10^ expected to be associated with changes in immunity. To investigate our hypotheses, we defined two population models within our cohort. Model 1 was based on HSV-2 infection status at the study visits grouped into: 1) HSV-2 negative, 2) incident HSV-2, and 3) established HSV-2. Model 2 was defined by HSV-2 acquisition status as: 1) remaining negative throughout the study with a minimum of two HSV-seronegative visits, and 2) seroconverted during the study. The median number of visits for women remaining HSV-2 negative was 2, and the median number of visits for women with incident and established HSV-2 infections was 3.

To ensure each infection at each visit could be accurately categorized as incident or established by the above criteria, we excluded: 1) seropositive baseline visits or visits <6 months from baseline as in both cases we could not be certain that seroconversions occurred before or after 6 months. 2) HSV-2 seronegative visits less than 3 months before seroconversion because the subject might have been already infected at that time and not yet developed antibodies, and 3) HSV-2 negative visits followed by exit from the study within 3 months. due to lack of serology test at the exit visit. We identified 3116 visits from 1019 women (413 Ugandan and 606 Zimbabwean) that met our inclusion criteria and infection definitions.

### Laboratory Diagnosis of Infection

HSV-2 status was determined by a type-specific serological IgG antibody enzyme-linked immunosorbent assay (ELISA) (Focus Technologies, Cypress, California, USA) as previously described^3^. *C. trachomatis* (CT) and *N. gonorrhoeae* (NG) were diagnosed by PCR, *T. vaginalis* (TV) and *Candida* by wet mount. Abnormal microbiota and bacterial vaginosis (BV) were assessed by Nugent scoring. HIV status was determined by ELISA and confirmed by PCR.

### Measurement of Biomarkers

Cervical swabs were collected and analyzed for immune biomarkers as previously described^11^. Ten biomarkers were chosen for their proven role in vaginal innate immunity and reliable detection in cervical secretions^11-13^. Interleukin (IL)-1β, IL-6, IL-8, IL-1 receptor antagonist (IL-1RA), RANTES, MIP-3α, VEGF, and sICAM-1 were measured in cervical secretions by a multiplex (Meso Scale Discovery (MSD), Gaithersburg, MD, USA). Secretory Leukocyte Peptidase Inhibitor (SLPI) was measured by R&D Systems ELISA (Minneapolis, MN, USA) and Beta-Defensin-2 (BD-2) – by Phoenix Pharmaceuticals ELISA (Burlingame, CA, USA). Serum IL-6 and IL-7 were measured by an MSD multiplex and serum C-Reactive Protein (CRP) and soluble (s)CD14 by a Luminex multiplex (Bio-Techne R&D Systems). Duplicate measurements of each specific protein were averaged and normalized to average total protein (Pierce BCA assay, Fisher Scientific, Pittsburgh, PA, USA).

### Statistical Analyses

We compared participants’ baseline characteristics by HSV-2 status (HSV-2 negative visits, HSV-2 incident visits and HSV-2 established visits) using joint chi-square tests via the generalized estimating equation approach and the Freeman-Halton test if numbers of visits were less than 5. Participants providing at least one sample for analysis and participants with both unpaired and paired samples (had both cervical and serum biomarkers) were included. Because immunity biomarker levels do not follow Gaussian distribution, concentrations were normalized using Box-Cox power transformation. Serum samples were analyzed in one batch. Cervical biospecimens were analyzed in two assay batches 4 years apart and data were harmonized for batch variation as previously described^7^.

We applied generalized linear mixed-effects models to evaluate if levels of systemic and cervical immune mediators differ among HSV-2 negative, HSV-2 incident, or HSV-2 established visits and adjusted for covariates. We performed bivariate analysis to determine the estimated odds ratio of HSV-2 seroconversion with individual/grouped biomarker levels activated/suppressed in HSV-2 seroconverters at the quarterly visit prior to the incident visit and negative visits from individuals remaining HSV-2 negative. The grouped analysis replicated biomarker combinations previously examined as predictors of HIV-1 acquisition (reference). P-values less than 0.05 were considered statistically significant. Statistical analyses were performed using SAS 9.4 (SAS Institute, Cary, NC, USA).

## Results

### Cohort Demographics & Descriptive Statistics

This analysis included data from 3116 HIV-1 negative visits from Zimbabwe (64%) and Uganda (36%) of which 1505 were HSV-2 negative (48.3%), 633 were HSV-2 incident (20.3%) and 978 were HSV-2 established (31.4%) visits (Table 1). Most visits (59%) were from women 18-24 years old. Visits were equally distributed by DMPA, COCs and no-hormonal method use. Few visits (8%) were from pregnant and from breastfeeding women (15%). Almost a third of visits (28%) were from women with BV and 11% had candidiasis while chlamydia (2%), gonorrhea (2%) and trichomoniasis (3%) were rare.

**Table 1.**
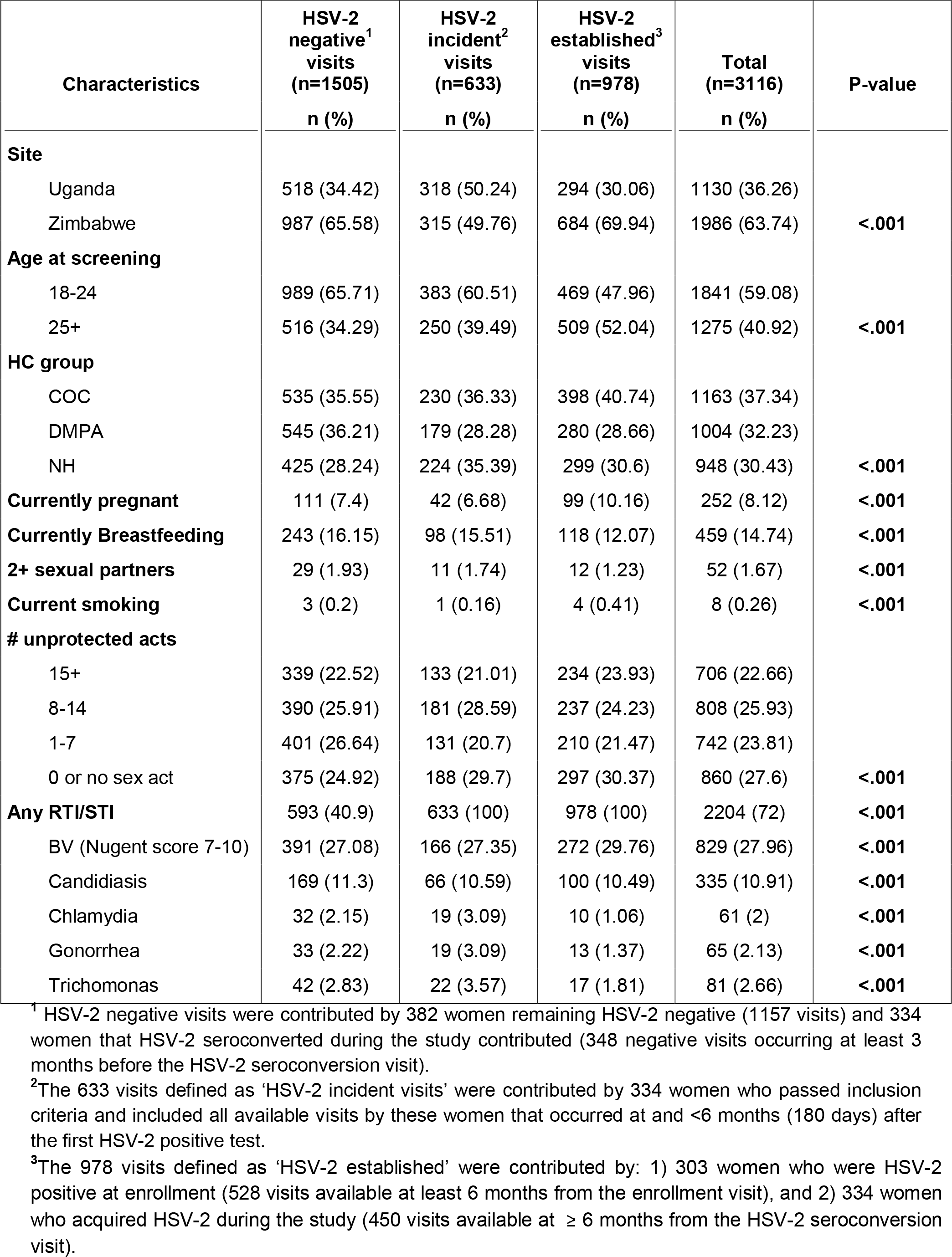
Characteristics of participant visits by HSV-2 status among HIV negative visits

HSV-2 negative visits were more likely from Zimbabwe (66%) and from younger women (66%). The majority of HSV-2 negative visits were contributed by women who remained HSV-2 negative (77%) while the remainder were collected from HSV-2 seroconverters at least 3 months prior to the incident visit. Incident visits were more likely from younger women (61%) while established visits were more likely from older women (52%), from women from Zimbabwe (70%) and women with BV (30%) (Table 1).

### Cervical and Systemic Biomarkers Preceding and Predicting HSV-2 Acquisition

To determine whether systemic immunity may contribute to the risk of HSV-2 acquisition, either independently or in conjunction with altered cervical immunity, we first measured individual cervical and systemic biomarkers at the 3-month visit preceding HSV-2 seroconversion. Then we assessed whether inflammatory or immunosuppressive status concomitant at both systemic and cervical sites predisposes to HSV-2.

#### Individually Altered Biomarkers (Table 2)

In bivariate modeling we found that 5 of the 10 cervical and 2 of the 4 systemic biomarkers were individually associated with subsequent HSV-2 acquisition. Higher odds were found with cervical high BD-2 (OR=1.45, 95% CI 1.09-1.93, P=0.01), low SLPI (OR=1.50, 95% CI 1.13-2.00, P<0.01) or low ICAM-1 (OR=1.41, 95% CI 1.06-1.88, P=0.02). Lower odds of HSV-2 acquisition were found with cervical high IL-6 or high MIP-3α (OR=0.75, 95% CI 0.56-1.00, P<0.05). Systemic markers associated with subsequent HSV-2 acquisition included high sCD14 (OR=1.93, 95% CI 1.34-2.78, P<0.001) and IL-6 (OR=1.53, 95% CI 1.09-2.14, P=0.01).

**Table 2.** Estimated odds ratios (OR and 95% confidence intervals) for acquiring HSV-2 within the next 3 months with levels of individual and grouped biomarkers measured at the HSV-2 negative visit preceding serocoversion and compared to the visits contributed by the control women who never acquired HSV-2.

| Category | Biomarker | Samples ↑ or ↓<sup>1</sup> / Sample used<sup>2</sup> | OR<sup>3</sup> (95% CI) | P-value |
| --- | --- | --- | --- | --- |
| **Cervical** | BD2↑ | 677 / 1365 | **1.45 (1.09, 1.93)** | **0.012** |
|  | SLPI↓ | 661 / 1366 | **1.50 (1.13, 2.00)** | **0.006** |
|  | IL-1RA↓ | 669 / 1366 | 1.18 (0.89, 1.57) | 0.256 |
|  | IL-1β↑ | 697 / 1366 | 0.83 (0.62, 1.10) | 0.200 |
|  | IL-6↑ | 696 / 1366 | **0.75 (0.56, 1.00)** | **0.048** |
|  | IL-8↑ | 716 / 1366 | 0.78 (0.58, 1.03) | 0.082 |
|  | MIP-3α↑ | 696 / 1366 | **0.75 (0.56, 1.00)** | **0.048** |
|  | RANTES↑ | 690 / 1366 | 1.07 (0.81, 1.43) | 0.628 |
|  | ICAM-1↓ | 672 / 1366 | **1.41 (1.06, 1.88)** | **0.018** |
|  | VEGF↓ | 648 / 1366 | 1.07 (0.81, 1.43) | 0.630 |
|  | IL-1β↑, IL-6↑ | 509 / 1366 | **0.58 (0.42, 0.80)** | **0.001** |
|  | IL-8↑, MIP-3α↑ | 535 / 1366 | **0.68 (0.50, 0.92)** | **0.012** |
|  | ICAM-1↓, VEGF↓ | 366 / 1366 | **1.47 (1.08, 2.00)** | **0.013** |
| **Serum** | sCD14↑ | 192 / 787 | **1.93 (1.34, 2.78)** | **<0.001** |
|  | CRP↓ | 391 / 787 | 1.14 (0.82, 1.59) | 0.439 |
|  | IL-6↑ | 391 / 787 | **1.53 (1.09, 2.14)** | **0.014** |
|  | IL-7↑ | 392 / 787 | 1.17 (0.84, 1.63) | 0.366 |
|  | IL-6↑, IL-7↑ | 231 / 787 | 1.32 (0.92, 1.88) | 0.130 |
|  | sCD14↑, IL-6↑ | 130 / 787 | **2.23 (1.49, 3.35)** | **<0.001** |
|  | sCD14↑, IL-6↑, IL-7↑ | 80 / 787 | **1.73 (1.05, 2.86)** | **0.033** |

<sup>1</sup> ↑ indicates levels above and ↓ below median of all 1505 HSV-2 negative study visits for all biomarkers except sCD14 where ↑ indicates levels above top quartile.
<sup>3</sup>Odds ratios (OR) and 95% confidence intervals (CI) estimate likelihood of subsequent HSV-2 seroconversion with biomarker levels at the HSV-2 negative visit either below or above median/top quartile.

#### Concomitantly Altered Biomarkers within Each Anatomic Compartment (Table 3)

We combined biomarkers within the same anatomic compartment to assess whether their concomitant activation or suppression was predictive of HSV-2 acquisition. In cervical secretions, concomitant high IL-1β and IL-6 (OR=0.58, 95% CI 0.42-0.80, P=0.001) or IL-8 and MIP-3α (OR=0.68, 95% CI 0.50-0.92, P=0.01) indicated decreased risk while low ICAM-1 and VEGF (OR=1.47, 95% CI 1.08-2.00, P=0.01) – increased risk of HSV-2 acquisition. Within the systemic circulation, increased HSV-2 acquisition risk was associated with the combinations of high sCD14 and IL-6 (OR=2.23, 95% CI 1.49-3.35, P<0.001) or high sCD14, IL-6, and IL-7 (OR=1.73, 95% CI 1.05-2.86, P=0.03).

**Table 3.** Differences in levels^1^ of cervical and systemic biomarkers by HSV-2 infection status adjusted for covariates

| Cervical Biomarker<sup>2</sup> | No. of specimens | HSV-2 status at the visit |  |  |
| --- | --- | --- | --- | --- |
|  |  | incident (n=491) vs. negative (n=1203) | established (n=838) vs. negative (n=1203) | incident (n=491) vs. established (n=838) |
| BD2 | 2530 |  | ↓ ** |  |
| SLPI | 2532 | ↓ * | ↓ ** |  |
| IL-1RA | 2532 | ↓ ** |  |  |
| IL-1β | 2532 | ↓ *** |  | ↓ ** |
| IL-6 | 2532 | ↓ *** | ↓ *** |  |
| IL-8 | 2532 | ↓ ** | ↓ * |  |
| MIP-3α | 2532 | ↓ *** |  | ↓ ** |
| RANTES | 2532 |  |  |  |
| ICAM-1 | 2532 |  |  |  |
| VEGF | 2532 | ↓ ** | ↓ * |  |
| Serum Biomarker<sup>3</sup> | No. of specimens | incident (n=390) vs. negative (n=646) | established (n=361) vs. negative (n=646) | incident (n=390) vs. established (n=361) |
| sCD14 | 1397 |  |  |  |
| CRP | 1397 |  |  |  |
| IL-6 | 1396 |  |  | ↓ ** |
| IL-7 | 1396 |  |  |  |

<sup>1</sup>↑ indicates significantly higher levels and ↓ indicates significant lower levels of these mediators compared to an HSV-2 negative reference (including all negative visits contributed by all women with information on adjusted variables listed in the models below) or the established HSV-2 reference \*\*\* P<0.001; \*\* P<0.01; \* P<0.05.
<sup>2</sup>The generalized linear mixed-effects models of differences by cervical biomarkers are adjusted for country, age, 5-level hormonal variable (pregnant/breastfeeding/majority COC/majority DMPA/majority NH), unprotected sex acts, current sexually transmitted or reproductive tract infections (STI/RTI), vaginal drying and cleaning practices.
<sup>3</sup>The generalized linear mixed-effects models of differences by serum biomarkers are adjusted for country, age, 5-level hormonal variable (pregnant/breastfeeding/majority COC/majority DMPA/majority NH), and current sexually transmitted or reproductive tract infections (STI/RTI).

#### *Combined Cervical and Systemic Biomarkers* (Figure 1 and Supplementary Table 1)

We found additional significant predictive patterns when we combined concomitantly aberrant cervical and systemic biomarkers. High systemic sCD14 in combination with either low cervical SLPI (OR=1.85, 95% CI 1.09-3.12, P=0.02) or high cervical BD-2 (OR=2.18, 95% CI 1.26-3.75, P=0.005) or low cervical VEGF and ICAM-1 (OR=2.02, 95% CI 1.04-3.90, P=0.04) conveyed higher odds of HSV-2 acquisition at the subsequent visit. High systemic IL-6 in combination with low cervical VEGF and ICAM-1 (OR=1.94, 95% CI 1.16-3.23, P=0.01) or with low cervical SLPI (OR=1.65, 95% CI 1.08-2.53, P=0.02) was associated with increased HSV-2 acquisition while high systemic IL-6 in combination with high cervical IL-1β and IL-6 was associated with decreased HSV-2 acquisition (OR=0.44, 95% CI 0.24-0.83, P=0.01). Low systemic CRP in combination with high cervical BD-2 (OR=1.82, 95% CI 1.20-2.76, P=0.005) was associated with increased HSV-2 acquisition.

**Figure 1.**
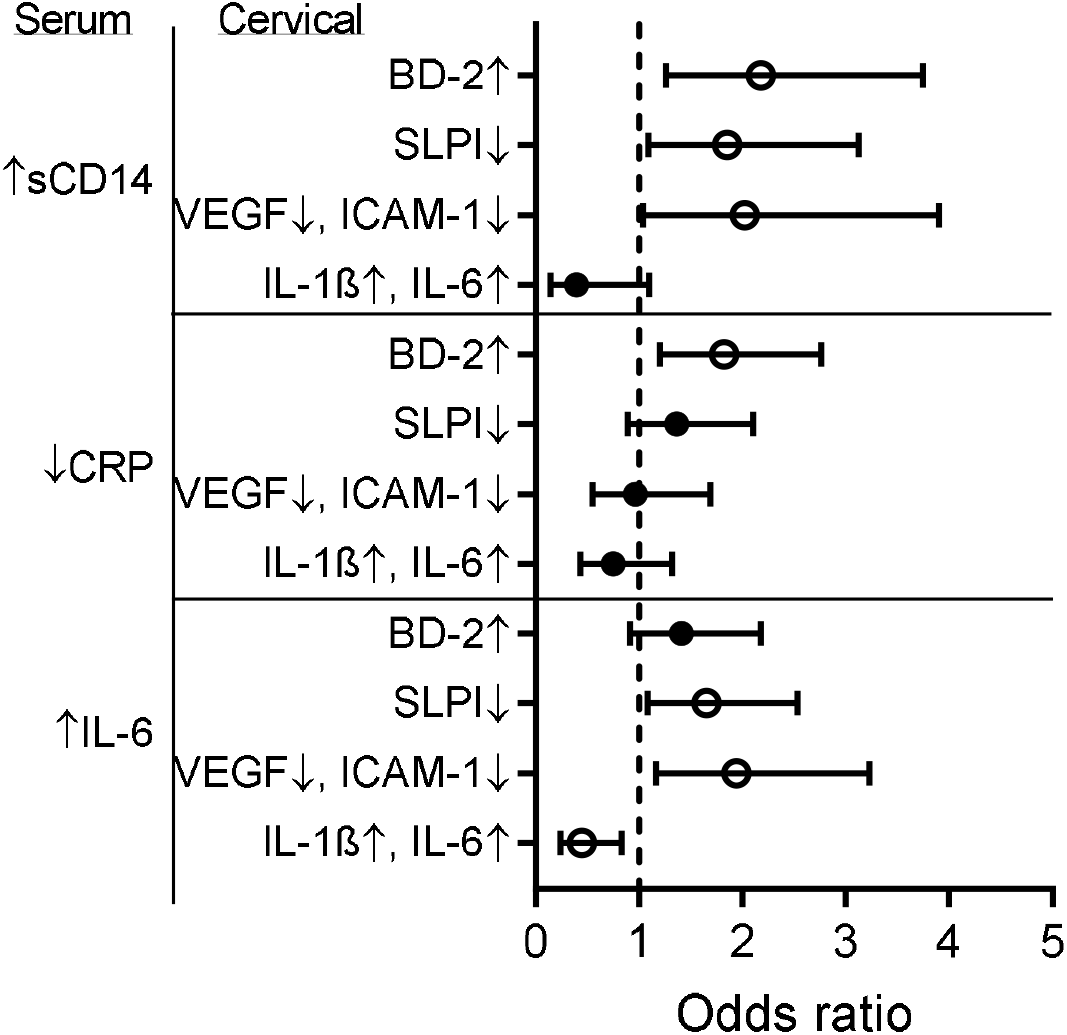
Forest plot of estimated odds ratios of HSV-2 acquisition for levels of concomitantly imbalanced systemic and cervical Immunity assessed 3 months prior to the HSV-2 seroconversion visit. The odds ratios with 95% confidence intervals are denoted by open circles for significant (p<0.05) and closed circles for non-significant p-values. ↑ indicates levels above and ↓ below the median of all HSV-2 negative study visits for all biomarkers except sCD14 where ↑ indicates levels above top quartile. Bivariate analysis compared HSV-2 seroconverters at the visit prior to the incident visit to HSV-2 negative visits from individuals remaining HSV-2 negative throughout.

### Differences in Cervical and Systemic Immunity by HSV-2 Incident and Established Infection Status

Analysis adjusted for relevant covariates confirmed our hypotheses that HSV-2 infection changes cervical and systemic innate immunity, and that incident and established infections differentially influence these changes. We found that HSV-2 incident visits had lower levels of 7/10 cervical immune biomarkers compared to all HSV-2 negative visits contributed by women throughout the study. Significant differences included lower SLPI, IL-1RA, IL-1β, IL-6, IL-8, MIP-3α and VEGF (Table 3). In contrast, HSV-2 established visits had lower levels of 5/10 cervical mediators including SLPI, IL-6, IL-8 and VEGF and adding significantly lower BD-2 (p<0.01) compared to the HSV-2 negative visits. In a direct comparison of incident vs. established visits, we found that women with incident infections had lower levels of cervical IL-1β and MIP-3α, but also lower systemic IL-6 compared to women with established HSV-2 infections (p<0.01).

## Discussion

This study provides evidence that while imbalances in some cervical innate immunity mediators may precede and predict both HIV-1 and HSV-2 infection, the two viral infections can be distinguished by antecedent patterns derived from both the mucosal and peripheral immune compartments, suggesting both common and divergent mechanisms of antiviral defense. Moreover, we show that HSV-2 infection not only changes innate immunity parameters previously associated with HIV risk but that the changes occurring within 6 months of HSV-2 infection differ from those observed 6-15 months later. These differences shed light on potential mechanisms, underlying epidemiologic findings of a greater HIV acquisition risk with more recent HSV-2 infection.

To our knowledge, this study is the first to implicate systemic immunity and combined mucosal-systemic patterns in HSV-2 risk. We had previously shown that top quartile levels of cervical IL-6, SLPI and ICAM-1 decrease the likelihood of HSV-2 incidence^8^. Consistent with and expanding those findings we now report that a generalized immunosuppressive state in the cervical compartment, characterized by SLPI↓, IL-6↓ (alone or combined with IL-1β↓), MIP-3α↓ (alone or combined with IL-8↓), or ICAM-1↓ (alone or combined with VEGF), predicts incident HSV-2 infection. BD-2 was the only upregulated marker preceding HSV-2 acquisition. In contrast to the permissive immunosuppressed status at the cervix, the systemic circulation displayed immunoinflammatory activation (serum sCD14↑, IL-6↑, or concomitant increase of both) to be predictive of HSV-2 acquisition.

SLPI is an antimicrobial protein with anti-inflammatory properties^14^ and has been shown to inhibit both HSV-2^15^ and HIV-1^16^ infection, which is consistent with our results of cervical SLPI↓ being predictive of both HSV-2 (reported here) and HIV-1 acquisition (reported previously^7^). BD-2 is also an antimicrobial peptide and is part of the protective host responses to infection of the vaginal mucosa^17^. However, BD-2 also has chemotactic activity^17^ which may increase recruitment of HIV target cells to mucosal sites thereby facilitating viral transmission, which may explain why higher BD2 was associated with risk of HIV acquisition^7^. Other studies (reviewed in^18,19^) suggest that the role of both alpha and beta defensins in viral infection may be more complex *in vivo* and may be altered by other factors in the female genital tract such as the microbiome and STIs. It is possible that high cervical BD-2 is a consequence of an underlying undiagnosed microbiome shift or asymptomatic STI.

Systemic immunity can be functionally distinct from mucosal immunity in the female genital tract^20,21^ and measuring markers in both compartments has provided distinct biomarkers of HIV acquisition risk ^7,22^. Prior to our study there were no published data on systemic immunity preceding HSV-2 acquisition. We report that high systemic sCD14 and IL-6, both alone and combined, predict HSV-2 acquisition. Interestingly, the effect of higher sCD14 on HSV-2 risk was offset by concomitant higher cervical IL-6 and IL-1β. IL-6 is a pleiotropic cytokine implicated in the pathogenesis of various infectious and non-infectious diseases^23^. IL-6 is part of the antiviral stress response and thus its mucosal upregulation may be a sign of more robust anti-viral barrier. Targeting IL-6 systemically constitutes a treatment strategy for inflammation-mediated diseases including diabetes, obesity, and rheumatoid arthritis^24^. Systemically altered IL-6 may play a role in malnourishment pathophysiology^25^ and its levels in the circulation are also controlled by genetic polymorphisms. Therefore, it is possible that high systemic IL-6 may not directly affect HSV-2 pathogenesis at the mucosal site but could be indicative of an underlying unmeasured exposure, disease, or condition that may independently increase susceptibility to HSV-2. sCD14 is a marker for monocyte activation and microbial translocation from the mucosal surface to the systemic circulation and may be a sign of mucosal damage^26^. Bacterial endotoxins and inflammatory cytokines such as IL-6 can induce the release of sCD14 in the circulation^27^. Considering this biological link between IL-6 and sCD14, it is not unexpected that their serum levels show the same direction of association.

The combination of systemic CRP↓ and cervical BD-2↑, associated with HSV-2 risk in this study was also found to precede HIV-1 acquisition^7^. CRP is primarily produced in the liver but also by lymphocytes and other cell types and low CRP may interfere with its important part in innate antiviral immune responses including completement activation ^28^.

We have further demonstrated suppressed innate immunity with differences between women with incident and established HSV-2 infections. Women with incident infections had lower levels of 7 cervical markers (SLPI, IL-1RA, IL-1β, IL-6, IL-8, MIP-3α and VEGF) while women with established infections had lower levels of 4 of these 7 markers and lower cervical BD-2 compared to HSV-2 negative visits. In a direct comparison to established infections, women with incident infections had lower levels of cervical IL-1β and MIP-3α (an anti-HIV microbicide^29^) and lower systemic IL-6. The lower serum IL-6 previously found predictive of HIV-1 acquisition^7^ offered one plausible biological link between higher risk for HIV-1 with incident than established HSV-2 infections. The overall immunosuppressed state may also contribute to higher HIV susceptibility among both incident and prevalent infections compared to HSV-2 negative women.

Our study has several important strengths. We followed over a thousand women with roughly equal-sized groups of women with incident or established HSV-2 infections and women remaining HSV-2 negative. We were able to analyze large numbers of participant visits with matched cervical and serum specimens thus allowing us to examine the relative and combined contributions of immune factors from each of these anatomic compartments. HSV-2 infection was measured at each 12-week visit providing us with accurate information about the timing of incident infections and accurately identifying the three HSV-status groups. For the HSV-2 infection status model, we adjusted for relevant confounders thus strengthening associations between the biomarkers and incident vs. established infections. In the predictive model we decided not to control for behavioral and demographic factors as prior evidence^8^ from this cohort suggested that the immune markers were more proximate to HSV-2 infection and thus control for antecedent variables was inappropriate. Additionally, all biomarkers were measured at the same accredited laboratory with methods previously validated for technical accuracy and clinical content^11^. A limitation of the studied dataset is that not all women contributed both HSV incident and established visits, and thus we were unable to do incident-established comparisons where all women served as their own controls. Future validation studies should be conducted in ethnically and racially diverse populations and among larger groups of pregnant women.

## Conclusion

This study contributes to a better understanding of HSV-2 acquisition in women and the complex link between HSV-2 and HIV-1 by providing clinical evidence for shared biological mechanisms of vulnerability to viral infection based on alterations of systemic and mucosal innate immunity. By identifying molecular predictors of HSV-2 risk, this study provides targets and clinical safety endpoints for the development of preventive products.

## Data Availability

All data produced in the present study are available upon reasonable request to the corresponding author.

## Acknowledgements

The authors acknowledge the contributions of the study staffs and participants in Zimbabwe and Uganda. The authors thank the following members of the Fichorova Laboratory, who contributed to this study by processing the cervical samples and performing all protein assays (listed in alphabetical order): Bi Yu Li, Bisiayo Fashemi, Hassan Dawood, Hidemi Yamamoto, Huaiping Yuan, Noah Beatty, Olimpia Suciu, Raymond Wong, Ryan Murray, Tai Nguyen, Xenia Chepa-Lotrea, Yoshika Yamamoto, and Yujin Lee.

---

**Supplementary Table 1.** Estimated odds ratios (OR) of HSV-2 acquisition for levels of concomitantly imbalanced systemic and cervical Immunity measured at the HSV-2 negative visits from individuals remaining HSV-2 negative and the HSV-2 seroconverters at the visit prior to the incident visit.

| Systemic + Cervical Biomarker Groups | #↑↓<sup>1</sup> / #Sample used<sup>2</sup> | OR<sup>3</sup> (95% CI) | P-value |
| --- | --- | --- | --- |
| ***Concomitant serum sCD14 ↑ + cervical ↑↓*** |  |  |  |
| IL-1β↑, IL-6↑ | 48 / 703 | 0.39 (0.14, 1.11) | 0.077 |
| IL-8↑, MIP-3α↑ | 52 / 703 | 0.68 (0.30, 1.55) | 0.360 |
| VEGF↓, ICAM-1↓ | 47 / 703 | **2.02 (1.04, 3.90)** | **0.037** |
| SLPI↓ | 84 / 703 | **1.85 (1.09, 3.12)** | **0.022** |
| BD-2↑ | 72 / 702 | **2.18 (1.26, 3.75)** | **0.005** |
| IL-1RA↓ | 67 / 703 | 1.34 (0.72, 2.46) | 0.354 |
| RANTES↑ | 87 / 703 | 1.30 (0.75, 2.25) | 0.351 |
| ***Concomitant serum CRP ↓ + cervical ↑↓*** |  |  |  |
| IL-1β↑, IL-6↑ | 114 / 703 | 0.75 (0.43, 1.32) | 0.322 |
| IL-8↑, MIP-3α↑ | 125 / 703 | 1.15 (0.71, 1.88) | 0.568 |
| VEGF↓, ICAM-1↓ | 96 / 703 | 0.96 (0.55, 1.69) | 0.892 |
| SLPI↓ | 169 / 703 | 1.36 (0.89, 2.10) | 0.156 |
| BD-2↑ | 168 / 702 | **1.82 (1.20, 2.76)** | **0.005** |
| IL-1RA↓ | 179 / 703 | 1.18 (0.77, 1.82) | 0.446 |
| RANTES↑ | 207 / 703 | 1.21 (0.80, 1.83) | 0.357 |
| ***Concomitant serum IL6 ↑ + cervical ↑↓*** |  |  |  |
| IL-1β↑, IL-6↑ | 121 / 703 | **0.44 (0.24, 0.83)** | **0.012** |
| IL-8↑, MIP-3α↑ | 132 / 703 | 0.88 (0.53, 1.45) | 0.612 |
| VEGF↓, ICAM-1↓ | 89 / 703 | **1.94 (1.16, 3.23)** | **0.011** |
| SLPI↓ | 164 / 703 | **1.65 (1.08, 2.53)** | **0.020** |
| BD-2↑ | 156 / 702 | 1.41 (0.91, 2.18) | 0.125 |
| IL-1RA↓ | 156 / 703 | 1.41 (0.91, 2.19) | 0.123 |
| RANTES↑ | 174 / 703 | 1.07 (0.69, 1.66) | 0.766 |

<sup>1</sup> ↑ indicates activated (levels above median) and ↓ indicates suppressed (levels below median) immunity, with the exception of sCD14 which is activated if its concentration is above top quartile, for all HIV-1 and HSV-2 negative visits.
<sup>2</sup> HSV-2 negative visits from individuals remaining HSV-2 negative and HSV-2 seroconverters at the visit prior to the incident visit.
<sup>3</sup> Odds ratios (OR) and 95% confidence intervals (CI) estimated likelihood of HSV-2 seroconversion with biomarker levels activated/suppressed as indicated by arrows assessed concomitantly.

